# Alcohol consumption and long-term risk of gout in men and women: a prospective study addressing potential reverse causation

**DOI:** 10.1101/2024.02.28.24303525

**Authors:** Jie-Qiong Lyu, Xian-Zhen Peng, Jia-Min Wang, Meng-Yuan Miao, Hao-Wei Tao, Miao Zhao, Jie Zhu, Jing Yang, Jing-Si Chen, Li-Qiang Qin, Wei Chen, Guo-Chong Chen

## Abstract

**Background:** While specific alcoholic beverages have been associated with gout, the influence of residual confounding and potential reverse-causation bias on these associations remains to be addressed.

**Methods:** The exploratory analysis included 218,740 men and 271,389 women free of gout at recruitment of the UK Biobank. Among these, 181,925 men and 223,700 women remained for the final analysis where potential reverse causation was addressed, i.e., by excluding participants who had or were likely to have reduced alcohol intake due to health issues at baseline, in addition to cases that were identified within the first 2 years of follow-up.

**Results:** In the final analysis, current drinkers had a higher risk of gout than never drinkers in men (HR = 1.78, 95% CI: 1.39-2.28) but not in women (HR = 0.84, 95% CI: 0.68-1.03). Among current drinkers, higher alcohol consumption was associated a substantially higher risk of gout in men and a moderately higher risk in women. The most evident sex difference in the consumption of specific alcoholic beverages was observed for beer/cider (mean: 4.16 vs. 0.44 pints/week in men and women, respectively). Consumption of champagne/white wine, beer/cider, and spirits each was associated with a higher risk of gout in both sexes, with beer/cider showing the strongest association (HR _per 1 pint/d_ = 1.55, 95% CI: 1.49-1.61 in men; HR _per 1 pint/d_ = 1.71, 95% CI: 1.14-2.57 in women). In the exploratory analysis, low to moderate consumption of specific alcoholic beverages were widely associated with a lower risk of gout, whereas all these inverse associations were eliminated in the final analysis. For example, red wine intake was associated with a lower risk of gout in women in the exploratory analysis (HR _per 1 glass/d_ = 0.79, 95% CI: 0.69-0.90), but not after adjusting for other alcoholic beverages and addressing potential reverse causation (HR _per 1 glass/d_ = 0.91, 95% CI: 0.77-1.06).

**Conclusions:** Consumption of total and several specific alcoholic beverages is associated with a higher risk of gout in both sexes. The sex-specific associations for total alcohol consumption may be attributable to differences in the type of alcohol consumed rather than biological differences between men and women.

## Introduction

Gout, which forms in the presence of increased urate concentrations (1), is the most common inflammatory arthritis (2). The prevalence of gout is sex-specific and varies across the global regions, with a male-to-female ratio ranging from 2:1 to 4:1 in Europe and North America and a substantially higher ratio of approximately 8:1 in Asia (2).

A sustained elevation in serum urate levels is the main driver for the development of gout (2). Alcohol intake has been associated with elevated serum urate levels (3), and thus might eventually cause gout through hyperuricemia (4). A number of epidemiological studies have suggested total or specific alcohol intake to be associated with a higher risk of gout (4-13). Of note, these studies are limited by using a cross-sectional/case-control design (6, 8, 12, 13), or assessing the association in men only (4, 7, 8, 11). In a cohort study comprising 1,951 men and 2,476 women, consumption of beer or spirits was associated with a higher risk of gout more strongly in women than in men (5), indicating possible sex-specific associations.

Furthermore, the previous studies of alcohol consumption and incident gout have commonly used non-drinkers as the referent population, such that the impact of reverse causation on the examined association remains an open issue. It has been found that, when evaluating the connection between lifestyle factors and health outcomes, reverse causation can influence the magnitude and sometimes the direction of the examined association (14). In the case of alcohol consumption, individuals with illness or ill health may have abstained from alcohol, shifting into non-drinking or occasional-drinking categories (15), which may attenuate or reverse a positive association, or exaggerate an inverse association between alcohol consumption and health risk. Such reverse-causation bias has been acknowledged as a contributor to the widely observed U- or J-shaped relationship between alcohol consumption and risk of cardiovascular disease or premature mortality (15, 16).

To address these research gaps and to gain more accurate risk estimates, we conducted sex-specific analyses to investigate the associations of total and specific alcohol consumption with the long-term risk of gout, with a particular emphasis on addressing the potential reverse-causation bias.

## Methods

### Study design and population

The UK Biobank constitutes a vast prospective cohort investigation aiming to explore the fundamental determinants of diverse chronic conditions, encompassing genetic, environmental, and lifestyle factors (17). A comprehensive study protocol is accessible online (https://www.ukbiobank.ac.uk/media/gnkeyh2q/study-rationale.pdf.). Briefly, the study recruited around 500,000 men and women, aged between 37 and 73 years, during the period from 2006 to 2010. Enrolled participants were invited to visit one of the 22 assessment centers situated across England, Wales, and Scotland, where they underwent extensive questionnaire surveys, brief interviews, and physical assessments. The UK Biobank study received approval from the research ethics committee (REC reference for UK Biobank 11/NW/0382) and all participants provided informed consent at enrollment.

### Assessment of gout

In line with the previous analyses of data from UK Biobank (18-20), participants with gout at baseline were identified based on self-reported data, including a self-reported history of gout, use of urate-lowering therapy (allopurinol, probenecid, and/or sulfinpyrazone) or colchicine, in addition to a hospital diagnosis of gout. Incident gout during follow-up was determined from hospital diagnoses recorded in primary or secondary inpatient records, using the International Classification of Diseases version 10 (ICD-10) codes M10.0, M10.2, M10.3, M10.4, and M10.9.

### Assessment of alcoholic beverages

At recruitment, information on alcohol consumption was obtained using a computer-assisted touchscreen system. Participants were asked to report the status of alcohol drinking as never, previous, or current drinking. Current drinkers were asked to respond to additional questions regarding how much of each alcoholic beverage (red wine, champagne/white wine, beer/cider, spirits, and fortified wine) they consumed in an average week (or month for those who drank less than once per week) (21). We categorized weekly alcohol consumption into four categories: <1, 1-2, 3-4, and ≥5 times/week. Specific alcohol consumption was categorized into five groups as 0, >0-1, 2-3, 4-6, and ≥7 glasses of red wine, glasses of champagne/white wine, pints of beer/cider, measures of spirits, or glasses of fortified wine per week.

### Assessment of covariates

Information on sociodemographic factors, medical histories, and lifestyle behaviors was collected at baseline by nurse-led interviews and touchscreen questionnaires. Anthropometric and blood-pressure measurements were performed by trained staff following standard procedures. The Townsend deprivation index (TDI) was derived by combining 4 census variables (unemployment, non-car ownership, non-home ownership, and household overcrowding). Body mass index (BMI) was calculated as measured weight divided by the square of measured height (kg/m^2^). Baseline physical activity was assessed using the self-reported short-form International Physical Activity Questionnaire with data summarized and reported in MET minutes per week. A healthy diet score was calculated based on the consumption of 6 commonly eaten food groups: fruit and vegetables, whole grains, refined grains, red meat, processed meat, and fish (22). Diabetes was defined as a self-reported physician’s diagnosis or antidiabetic medication use, or an HbA1c level of ≥6.5%. Hypertension was determined by systolic blood pressure ≥140 mm Hg, diastolic blood pressure ≥90 mm Hg, or self-reported physician’s diagnosis or use of antihypertensive medications. Dyslipidemia was defined as a reported physician’s diagnosis or use of lipid-lowering medications.

### Statistical analysis

Participants with gout at baseline (n = 10,661) or with missing data on alcohol intake (n = 1621) were excluded, leaving 490,129 participants (218,740 men and 271,389 women) for the exploratory analysis. To mitigate the influence of potential reverse causation, the final analysis excluded the following participants: 1) participants who had reduced alcohol intake for illness/ill health; 2) participants self-rated as having poor health; 3) participants who had major CVD/cancer at baseline; and 4) participants who developed gout within the first 2 years of follow-up. There were 181,925 men and 223,700 women remained for the final analysis (**Supplementary Figure 1**).

Baseline participant characteristics were summarized based on status (for all participants) or frequency (for current drinkers) of total alcohol drinking in men and women. Data were reported as number, percentage, or mean ± SD where appropriate. Correlations among consumption of the specific alcoholic beverages were assessed using Spearman’s partial correlation coefficients.

Cox proportional hazards regression models were used to estimate sex-specific hazard ratios (HRs) and 95% confidence intervals (CIs) for the association of total or specific alcohol consumption with incident gout. Follow-up time was calculated from the date of enrollment through the date of diagnosis of gout, death, withdrawal from the study, or the end of the most recent follow-up for the approved dataset, whichever occurred first. Different multivariable models with an increasing degree of covariate adjustment were constructed. Model 1 was adjusted for age (y), ethnic background (White, Asian/Asian British, Black/Black British, mixed), TDI, smoking (never, former, current [<10, 10-<50, >50 pack-years]), total physical activity (MET-h/week), healthy-diet score, hypertension (yes, no), dyslipidemia (yes, no), and diabetes (yes, no). Model 2 was further adjusted for BMI (kg/m^2^). When evaluating the association between consumption of specific alcoholic beverages and incident gout, we used an additional model (model 3) where all other types of alcoholic beverages were concurrently added to the model 2. We also modelled the intake as a continuous variable and estimated the HR (95% CI) of gout for each additional 1-unit increment of a specific alcoholic beverage.

To address the influence of potential reverse-causation bias, the final analysis using the full model were repeated after excluding the aforementioned 4 groups of participants, both one by one and concurrently. All statistical tests were two-sided and the analyses were performed using Stata software (version 15.1; StataCorp).

## Results

### Baseline participant characteristics

**Table 1** presents baseline characteristics of the participants according to alcohol drinking status for men and women who were included in the exploratory analysis. In men, 2.87%, 3.57%, and 93.56% participants were never, former, and current drinkers, respectively, and the corresponding proportions in women were 5.86%, 3.64%, and 90.50%, respectively. Regardless of sex, compared with never drinkers, current drinkers were more likely to be ethnically white, were less likely to be never smokers or have dyslipidemia or diabetes, and had lower levels of TDI (higher socioeconomic status) and BMI. In men, current drinkers were older and more likely to have hypertension than never drinkers, whereas female current drinkers were younger and less likely to have hypertension than female never drinkers. Sex-specific participant characteristics according to the frequency of any alcohol intake among current drinkers are reported in **Supplementary Table 1**.

**Table 1.**
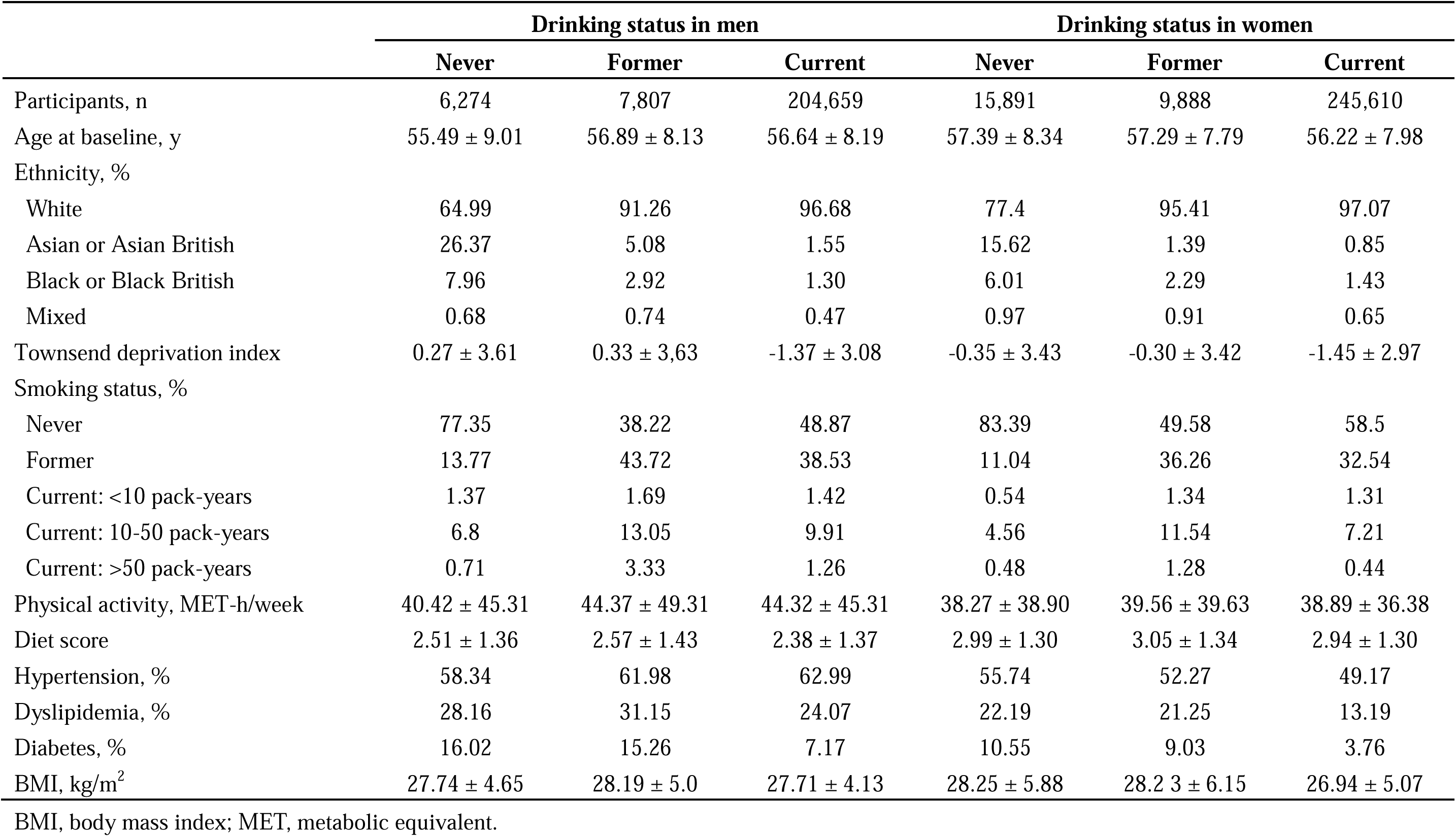
Baseline participants characteristics according to alcohol drinking status in men and women.

### Alcohol consumption and risk of gout in men and women

Among the participants in the exploratory analysis, 9,161 incident cases of gout (7,021 in men and 2,140 in women) were identified over a median follow-up time of 12.67 years. The number of cases was 5,804 (4,506 in men and 1,298 in women) among the participants included in the final analysis addressing potential reverse causation.

In men, as compared with never drinkers, current drinkers had a higher risk of gout in the exploratory analysis with the multivariable adjustment (model 2, HR = 1.58, 95% CI: 1.32-1.89), and the association was stronger in the final analysis accounting for potential reverse causation (HR = 1.78, 95% CI: 1.39-2.28) (**Table 2**). In women, after multivariable adjustment without the adjustment for BMI (model 1), current drinkers showed a lower risk of gout (HR = 0.78, 95% CI: 0.67-0.91) in the exploratory analysis, but the association was attenuated to be nonsignificant after further adjusting for BMI (model 2) and accounting for potential reverse causation in the final analysis (HR = 0.84, 95% CI: 0.68-1.03). Regardless of sex, the risk of gout did not differ between former and never drinkers across the analyses. A significant interaction between drinking status and sex was found (P for interaction <0.001 in the final analysis).

**Table 2.** Association between total alcohol consumption and gout in men and women.

|  | Never drinkers | Former drinkers | Current drinkers |
| --- | --- | --- | --- |
| **Men** |  |  |  |
| Case/Total | 132/6,274 | 203/7,807 | 6,686/204,659 |
| Model 1 | 1.00 (Referent) | 1.17 (0.94-1.46) | **1.51 (1.27-1.81)** |
| Model 2 | 1.00 (Referent) | 1.17 (0.93-1.46) | **1.58 (1.32-1.89)** |
| Model 2 <sup>a</sup> | 1.00 (Referent) | 1.16 (0.93-1.45) | **1.56 (1.31-1.87)** |
| Model 2 <sup>b</sup> | 1.00 (Referent) | 1.10 (0.85-1.41) | **1.62 (1.34-1.97)** |
| Model 2 <sup>c</sup> | 1.00 (Referent) | 1.24 (0.96-1.60) | **1.66 (1.35-2.04)** |
| Model 2 <sup>d</sup> | 1.00 (Referent) | 1.28 (1.00-1.63) | **1.66 (1.36-2.02)** |
| Model 2 <sup>e</sup> | 1.00 (Referent) | 1.23 (0.90-1.68) | **1.78 (1.39-2.28)** |
| **Women** |  |  |  |
| Case/Total | 199/15,891 | 117/9,888 | 1,824 /245,610 |
| Model 1 | 1.00 (Referent) | 0.98 (0.77-1.24) | **0.78 (0.67-0.91)** |
| Model 2 | 1.00 (Referent) | 0.95 (0.75-1.21) | 0.86 (0.73-1.01) |
| Model 2 <sup>a</sup> | 1.00 (Referent) | 0.96 (0.76-1.21) | **0.85 (0.73-0.99)** |
| Model 2 <sup>b</sup> | 1.00 (Referent) | 0.89 (0.68-1.16) | 0.89 (0.75-1.06) |
| Model 2 <sup>c</sup> | 1.00 (Referent) | 0.94 (0.71-1.23) | 0.87 (0.73-1.04) |
| Model 2 <sup>d</sup> | 1.00 (Referent) | 0.94 (0.73-1.21) | **0.84 (0.71-0.99)** |
| Model 2 <sup>e</sup> | 1.00 (Referent) | 0.93 (0.68-1.28) | 0.84 (0.68-1.03) |

Model 1: age (y), ethnic group (White, Asian/Asian British, Black/Black British, mixed), Townsend deprivation index, smoking (never, former, current [ $<10$ , $10-<50$ , $>50$ pack-years]), total physical activity (MET-h/week), diet habit, hypertension (yes, no), dyslipidemia (yes, no), and diabetes (yes, no).
Model 2: Model 1 + BMI ( $\text{kg/m}^2$ ).
<sup>a</sup> Excluding current drinkers who had reduced alcohol intake for illness or ill health at baseline (6,711 and 2,040 cases remained in men and women, respectively).
<sup>b</sup> Excluding participants self-rating as having poor health at baseline.
<sup>c</sup> Excluding participants with major cardiovascular disease or cancer at baseline.
<sup>d</sup> Excluding participants developing gout within the first 2 years of follow-up.
<sup>e</sup> Excluding all above.
P for interaction between men and women $<0.001$ in the final analysis.

We then evaluated the frequency of any alcohol consumption and risk of gout in current drinkers (**Table 3**). In male drinkers, the risk of gout increased with increasing alcohol intake, with an HR of 1.97 (95% CI: 1.78-2.19) in the final analysis comparing ≥5 with <1 time/week of alcohol consumption. In female drinkers, with the model-1 adjustment, more frequent consumption of alcohol was associated with a significantly lower risk of gout. Such an inverse association was reversed in the final analysis adjusting for BMI and accounting for reverse causation, with an HR of 1.29 (95% CI: 1.09-1.54) comparing ≥5 with <1 time/week of alcohol consumption.

**Table 3.** Association between total alcohol consumption and gout according to drinking frequencies among current drinkers in men and women.

|  | Frequency |  |  |  | P-trend |
| --- | --- | --- | --- | --- | --- |
|  | <1/week | 1-2/week | 3-4/week | ≥5/week |  |
| **Men** |  |  |  |  |  |
| Case/Total | 817/36,201 | 1,664/ 56,976 | 1,926/56,868 | 2,279/54,614 |  |
| Model 1 | 1.00 (Referent) | **1.32 (1.22-1.44)** | **1.51 (1.39-1.64)** | **1.72 (1.59-1.87)** | **<0.001** |
| Model 2 | 1.00 (Referent) | **1.38 (1.27-1.50)** | **1.65 (1.52-1.80)** | **1.97 (1.81-2.13)** | **<0.001** |
| Model 2 <sup>a</sup> | 1.00 (Referent) | **1.38 (1.26-1.51)** | **1.68 (1.54-1.83)** | **2.00 (1.83-2.17)** | **<0.001** |
| Model 2 <sup>b</sup> | 1.00 (Referent) | **1.41 (1.28-1.54)** | **1.68 (1.54-1.84)** | **2.01 (1.84-2.19)** | **<0.001** |
| Model 2 <sup>c</sup> | 1.00 (Referent) | **1.40 (1.28-1.54)** | **1.71 (1.56-1.88)** | **2.03 (1.85-2.23)** | **<0.001** |
| Model 2 <sup>d</sup> | 1.00 (Referent) | **1.34 (1.23-1.47)** | **1.58 (1.44-1.72)** | **1.91 (1.75-2.09)** | **<0.001** |
| Model 2 <sup>e</sup> | 1.00 (Referent) | **1.34 (1.20-1.50)** | **1.63 (1.47-1.82)** | **1.97 (1.78-2.19)** | **<0.001** |
| **Women** |  |  |  |  |  |
| Case/Total | 697/76,286 | 464/69,913 | 340/55,719 | 323/43,692 |  |
| Model 1 | 1.00 (Referent) | **0.85 (0.75-0.96)** | **0.82 (0.72-0.94)** | **0.86 (0.75-0.98)** | **0.030** |
| Model 2 | 1.00 (Referent) | 0.97 (0.86-1.09) | 1.02 (0.89-1.17) | 1.15 (1.00-1.32) | **0.038** |
| Model 2 <sup>a</sup> | 1.00 (Referent) | 0.98 (0.87-1.11) | 1.06 (0.92-1.21) | **1.20 (1.04-1.38)** | **0.006** |
| Model 2 <sup>b</sup> | 1.00 (Referent) | 1.01 (0.89-1.14) | 1.06 (0.92-1.22) | **1.20 (1.04-1.39)** | **0.010** |
| Model 2 <sup>c</sup> | 1.00 (Referent) | 0.94 (0.82-1.08) | 0.99 (0.85-1.15) | **1.19 (1.02-1.39)** | **0.021** |
| Model 2 <sup>d</sup> | 1.00 (Referent) | 0.98 (0.86-1.11) | 1.02 (0.88-1.18) | **1.16 (1.00-1.34)** | **0.045** |
| Model 2 <sup>e</sup> | 1.00 (Referent) | 1.00 (0.85-1.17) | 1.03 (0.87-1.22) | **1.29 (1.09-1.54)** | **0.004** |

Model 1: age (y), ethnic group (White, Asian/Asian British, Black/Black British, mixed), Townsend deprivation index, smoking (never, former, current [ $<10$ , $10-<50$ , $>50$ pack-years]), total physical activity (MET-h/week), diet habit, hypertension (yes, no), dyslipidemia (yes, no), and diabetes (yes, no).
Model 2: Model 1 + BMI ( $\text{kg/m}^2$ ).
<sup>a</sup> Excluding current drinkers who had reduced alcohol intake for illness or ill health at baseline.
<sup>b</sup> Excluding participants self-rating as having poor health at baseline.
<sup>c</sup> Excluding participants with major cardiovascular disease or cancer at baseline.
<sup>d</sup> Excluding cases occurring within the first 2 years of follow-up.
<sup>e</sup> Excluding all above.

### Specific alcoholic beverages and risk of gout in men and women

In current drinkers, correlations among the specific alcoholic beverages were weak, with the strongest correlations between red wine and champagne/white wine in men (r = 0.10), and between red wine and champagne/white wine in women (r = −0.07) (**Supplementary Figure 2**). As compared with women, men consumed slightly more red wine (mean: 3.51 vs. 2.47 glasses/week) and spirits (mean: 1.68 vs. 1.03 measures/week) but slightly less champagne/white wine (mean: 1.49 vs. 2.38 glasses/week) (**Figure 1**). In both sexes, the consumption of fortified wine was much lower than that of other alcoholic beverages (**Figure 1**). The most evident sex difference in the consumption was observed for beer/cider, with mean of 4.16 pints/week in men and 0.44 pints/week in women.

**Figure 1.**
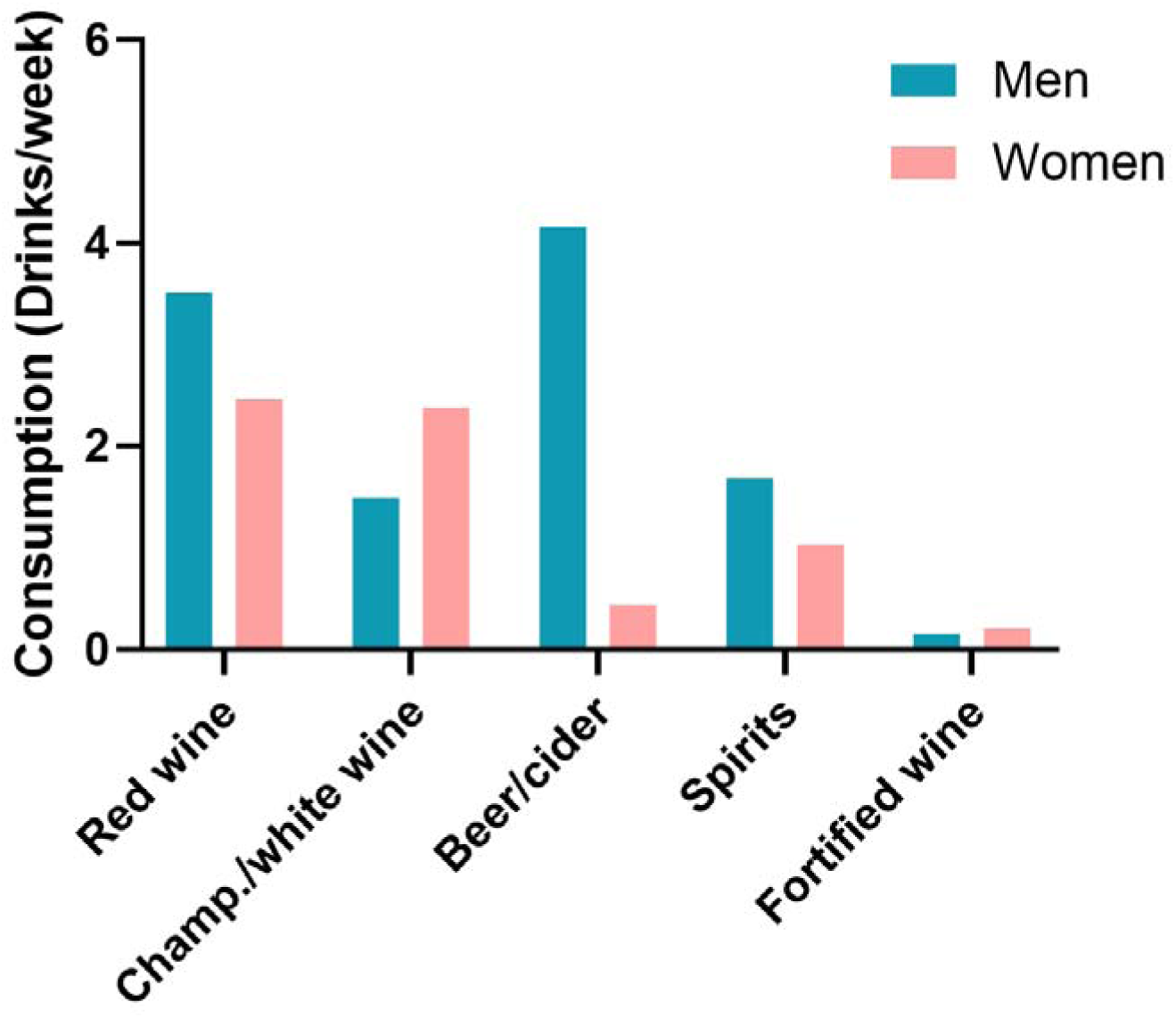
Average weekly consumption of alcoholic beverages.

We then investigated the relationships between different alcoholic beverages and risk of gout in men (**Table 4**) and women (**Table 5**). In the final analysis of current drinkers, consumption of champagne/white wine, beer/cider, and spirits each was associated with a higher risk of gout in both sexes, with beer/cider showing the strongest association (HR _per 1 pint/d_ = 1.55, 95% CI: 1.49-1.61 in men; HR _per 1 pint/d_ = 1.71, 95% CI: 1.14-2.57 in women). The association for spirits appeared to be stronger in women (HR _per 1 measure/d_ = 1.52, 95% CI: 1.30-1.78) than in men (HR _per 1 measure/d_ = 1.11, 95% CI: 1.05-1.16). Red wine was associated with a higher risk of gout only in men (HR _per 1 glass/d_ = 1.09, 95% CI: 1.04-1.14), and no significant associations were found for fortified wine either in men or in women.

**Table 4.** Association between consumption of specific alcoholic beverages and risk of gout among male current drinkers Weekly consumption of specific alcoholic beverages.

|  | Weekly consumption of specific alcoholic beverages |  |  |  |  | P-trend | Per 1<br>serving/day |
| --- | --- | --- | --- | --- | --- | --- | --- |
|  | 0/week | 0-<1/week | 2-3/week | 4-6/week | ≥7/week |  |  |
| *Red wine, glasses/week* |  |  |  |  |  |  |  |
| Case/Total | 2,270/61,695 | 492/17,720 | 1,033/33,014 | 1,005/32,881 | 1,037/28,902 |  |  |
| Model 1 | 1.00 (Referent) | **0.81 (0.73-0.89)** | **0.88 (0.82-0.95)** | **0.85 (0.79-0.92)** | 0.95 (0.88-1.02) | 0.147 | 1.00 (0.96-1.04) |
| Model 2 | 1.00 (Referent) | **0.84 (0.77-0.93)** | **0.93 (0.86-1.00)** | **0.90 (0.83-0.97)** | 1.01 (0.94-1.09) | 0.793 | 1.02 (0.98-1.07) |
| Model 3 | 1.00 (Referent) | 1.03 (0.92-1.15) | 1.07 (0.98-1.16) | 1.00 (0.92-1.09) | **1.15 (1.06-1.25)** | **0.010** | **1.08 (1.03-1.12)** |
| Model 3 <sup>a</sup> | 1.00 (Referent) | 0.99 (0.86-1.14) | 1.03 (0.93-1.14) | 1.00 (0.90-1.11) | **1.16 (1.05-1.29)** | **0.021** | **1.09 (1.04-1.14)** |
| *Champagne/white wine, glasses/week* |  |  |  |  |  |  |  |
| Case/Total | 3,526/99,971 | 624/22,411 | 875/29,117 | 567/16,721 | 329/8,376 |  |  |
| Model 1 | 1.00 (Referent) | **0.86 (0.79-0.93)** | **0.90 (0.83-0.97)** | 1.01 (0.92-1.10) | **1.14 (1.02-1.27)** | 0.121 | 1.09 (1.02-1.16) |
| Model 2 | 1.00 (Referent) | **0.88 (0.81-0.96)** | **0.93 (0.86-1.00)** | 1.04 (0.95-1.13) | **1.19 (1.05-1.32)** | **0.014** | **1.12 (1.05-1.19)** |
| Model 3 | 1.00 (Referent) | 1.04 (0.94-1.14) | 1.00 (0.92-1.08) | 1.10 (1.00-1.21) | **1.31 (1.16-1.48)** | **<0.001** | **1.17 (1.09-1.25)** |
| Model 3 <sup>a</sup> | 1.00 (Referent) | 1.10 (0.98-1.24) | 1.03 (0.93-1.14) | **1.14 (1.02-1.29)** | **1.48 (1.29-1.71)** | **<0.001** | **1.22 (1.13-1.32)** |
| *Beer/cider, pints/week* |  |  |  |  |  |  |  |
| Case/Total | 1,006/37,568 | 552/27,011 | 1,074/37,556 | 1,185/33,119 | 1,933/37,915 |  |  |
| Model 1 | 1.00 (Referent) | **0.83 (0.75-0.92)** | **1.16 (1.07-1.27)** | **1.47 (1.35-1.60)** | **2.05 (1.89-2.22)** | **<0.001** | **1.50 (1.45-1.55)** |
| Model 2 | 1.00 (Referent) | **0.83 (0.75-0.92)** | **1.15 (1.05-1.25)** | **1.42 (1.30-1.54)** | **1.97 (1.82-2.13)** | **<0.001** | **1.47 (1.42-1.52)** |
| Model 3 | 1.00 (Referent) | **0.86 (0.76-0.96)** | **1.18 (1.07-1.29)** | **1.49 (1.36-1.63)** | **2.13 (1.96-2.32)** | **<0.001** | **1.52 (1.47-1.58)** |
| Model 3 <sup>a</sup> | 1.00 (Referent) | 0.92 (0.80-1.05) | **1.18 (1.05-1.33)** | **1.47 (1.31-1.65)** | **2.21 (1.99-2.46)** | **<0.001** | **1.55 (1.49-1.61)** |
| *Spirits, measures/week* |  |  |  |  |  |  |  |
| Case/Total | 3,523/109,615 | 500/18,289 | 805/22,761 | 556/14,828 | 587/12,161 |  |  |
| Model 1 | 1.00 (Referent) | **0.84 (0.77-0.92)** | 1.02 (0.95-1.10) | 1.03 (0.94-1.13) | **1.24 (1.13-1.35)** | **<0.001** | **1.11 (1.07-1.16)** |
| Model 2 | 1.00 (Referent) | **0.83 (0.76-0.91)** | 0.99 (0.92-1.07) | 0.98 (0.89-1.07) | **1.15 (1.06-1.26)** | **0.005** | **1.08 (1.03-1.12)** |
| Model 3 | 1.00 (Referent) | 0.95 (0.86-1.05) | 1.02 (0.94-1.11) | 0.95 (0.86-1.04) | **1.17 (1.06-1.29)** | **0.024** | **1.09 (1.04-1.14)** |
| Model 3 <sup>a</sup> | 1.00 (Referent) | 1.00 (0.89-1.13) | 1.04 (0.94-1.15) | 0.97 (0.86-1.09) | **1.28 (1.13-1.43)** | **0.003** | **1.11 (1.05-1.16)** |
| *Fortified wine, glasses/week* |  |  |  |  |  |  |  |
| Case/Total | 5,616/165,253 | 236/8,505 | 140/4,434 | 64/1,434 | 12/389 |  |  |
| Model 1 | 1.00 (Referent) | **0.83 (0.73-0.95)** | 0.88 (0.74-1.04) | 1.20 (0.94-1.54) | 0.76 (0.43-1.34) | 0.506 | 0.87 (0.67-1.13) |
| Model 2 | 1.00 (Referent) | **0.85 (0.74-0.96)** | 0.89 (0.75-1.05) | 1.23 (0.96-1.57) | 0.83 (0.47-1.47) | 0.749 | 0.92 (0.71-1.19) |
| Model 3 | 1.00 (Referent) | 0.96 (0.83-1.10) | 0.92 (0.77-1.11) | **1.36 (1.05-1.76)** | 0.88 (0.47-1.64) | 0.504 | 1.04 (0.80-1.36) |
| Model 3 <sup>a</sup> | 1.00 (Referent) | 1.02 (0.86-1.20) | 0.97 (0.79-1.21) | 1.33 (0.96-1.83) | 1.37 (0.74-2.56) | 0.157 | 1.21 (0.84-1.54) |

Model 1: age (y), ethnic group (White, Asian/Asian British, Black/Black British, mixed), Townsend deprivation index, smoking (never, former, current [ $<10$ , $10-<50$ , $>50$ pack-years]), total physical activity (MET-h/week), diet habit, hypertension (yes, no), dyslipidemia (yes, no), and diabetes (yes, no).
Model 2: Model 1 + BMI ( $\text{kg/m}^2$ ).
Model 3: Model 2 + other types of alcoholic beverages.
<sup>a</sup> Excluding participants who had reduced alcohol intake for illness or ill health, participants self-rating as having poor health, and participants with major cardiovascular disease or cancer at baseline, in addition to cases occurring within the first 2 years of follow-up.

**Table 5.** Association between consumption of specific alcoholic beverages and risk of gout among female current drinkers Average weekly specific alcohol consumption.

|  | Average weekly specific alcohol consumption |  |  |  |  | P-trend | Per 1<br>serving/day |
| --- | --- | --- | --- | --- | --- | --- | --- |
|  | 0/week | 0-<1/week | 2-3/week | 4-6/week | ≥7/week |  |  |
| *Red wine, glasses/week* |  |  |  |  |  |  |  |
| Case/Total | 642/75,089 | 154/23,874 | 228/38,028 | 185/31,921 | 99/9,090 |  |  |
| Model 1 | 1.00 (Referent) | **0.80 (0.67-0.95)** | **0.76 (0.66-0.89)** | **0.74 (0.63-0.87)** | **0.63 (0.51-0.78)** | **<0.001** | **0.71 (0.62-0.81)** |
| Model 2 | 1.00 (Referent) | **0.83 (0.69-0.99)** | **0.83 (0.72-0.97)** | **0.82 (0.69-0.97)** | **0.72 (0.58-0.90)** | **0.001** | **0.79 (0.69-0.90)** |
| Model 3 | 1.00 (Referent) | 0.94 (0.77-1.15) | 0.92 (0.78-1.08) | 0.88 (0.74-1.06) | **0.77 (0.62-0.97)** | **0.030** | **0.83 (0.72-0.96)** |
| Model 3 <sup>a</sup> | 1.00 (Referent) | 0.78 (0.59-1.02) | 1.00 (0.81-1.22) | 0.93 (0.75-1.15) | 0.82 (0.63-1.08) | 0.427 | 0.91 (0.77-1.06) |
| *Champagne/white wine, glasses/week* |  |  |  |  |  |  |  |
| Case/Total | 562/72,919 | 177/29,686 | 245/38,178 | 166/29,019 | 122/17,880 |  |  |
| Model 1 | 1.00 (Referent) | **0.82 (0.70-0.98)** | 0.94 (0.81-1.09) | 0.88 (0.74-1.04) | 1.01 (0.83-1.23) | 0.874 | 0.99 (0.87-1.12) |
| Model 2 | 1.00 (Referent) | **0.82 (0.69-0.97)** | 0.97 (0.84-1.13) | 0.94 (0.79-1.11) | 1.15 (0.95-1.41) | 0.206 | 1.08 (0.95-1.22) |
| Model 3 | 1.00 (Referent) | 0.91 (0.76-1.10) | 1.01 (0.86-1.19) | 0.97 (0.80-1.16) | 1.21 (0.98-1.49) | 0.102 | 1.11 (0.97-1.26) |
| Model 3 <sup>a</sup> | 1.00 (Referent) | 0.89 (0.70-1.14) | 0.99 (0.80-1.21) | 1.03 (0.83-1.30) | **1.32 (1.03-1.71)** | **0.019** | **1.17 (1.01-1.35)** |
| *Beer/cider, pints/week* |  |  |  |  |  |  |  |
| Case/Total | 1,055/149,704 | 133/24,152 | 81/12,797 | 45/5,465 | 15/1,105 |  |  |
| Model 1 | 1.00 (Referent) | 0.92 (0.77-1.11) | 1.07 (0.85-1.35) | 1.32 (0.98-1.79) | 2.09 (1.25-3.50) | **0.004** | **1.56 (1.14-2.13)** |
| Model 2 | 1.00 (Referent) | 0.90 (0.75-1.08) | 1.11 (0.89-1.40) | 1.32 (0.98-1.79) | **2.67 (1.36-3.78)** | **0.001** | **1.64 (1.20-2.24)** |
| Model 3 | 1.00 (Referent) | 0.95 (0.78-1.15) | 1.09 (0.85-1.39) | **1.42 (1.04-1.94)** | **2.21 (1.27-3.83)** | **0.001** | **1.79 (1.29-2.49)** |
| Model 3 <sup>a</sup> | 1.00 (Referent) | 0.82 (0.63-1.07) | 1.10 (0.81-1.50) | 1.28 (0.84-1.95) | **2.59 (1.33-5.05)** | **0.009** | **1.71 (1.14-2.57)** |
| *Spirits, measures/week* |  |  |  |  |  |  |  |
| Case/Total | 768/129,541 | 134/20,904 | 172/21,302 | 132/12,259 | 93/7,216 |  |  |
| Model 1 | 1.00 (Referent) | 1.04 (0.86-1.25) | **1.26 (1.07-1.49)** | **1.58 (1.31-1.90)** | **1.70 (1.36-2.11)** | **<0.001** | **1.52 (1.34-1.72)** |
| Model 2 | 1.00 (Referent) | 0.96 (0.80-1.16) | 1.18 (1.00-1.39) | **1.46 (1.21-1.76)** | **1.56 (1.26-1.94)** | **<0.001** | **1.43 (1.26-1.62)** |
| Model 3 | 1.00 (Referent) | 1.04 (0.85-1.27) | **1.21 (1.01-1.44)** | **1.38 (1.12-1.68)** | **1.59 (1.26-2.00)** | **<0.001** | **1.46 (1.27-1.66)** |
| Model 3 <sup>a</sup> | 1.00 (Referent) | 1.19 (0.93-1.52) | **1.30 (1.04-1.61)** | **1.43(1.11-1.85)** | **1.71 (1.28-2.28)** | **<0.001** | **1.52 (1.30-1.78)** |
| *Fortified wine, glasses/week* |  |  |  |  |  |  |  |

| Case/Total | 1,191/173,472 | 63/11,960 | 67/6,530 | 15/2,270 | 8/588 |  |  |
| --- | --- | --- | --- | --- | --- | --- | --- |
| Model 1 | 1.00 (Referent) | **0.70 (0.54-0.90)** | **1.28 (1.00-1.64)** | 0.76 (0.45-1.26) | 1.39 (0.69-2.78) | 0.532 | 1.01 (0.66-1.56) |
| Model 2 | 1.00 (Referent) | **0.70 (0.55-0.91)** | **1.30 (1.02-1.67)** | 0.80 (0.48-1.33) | 1.56 (0.77-3.12) | 0.319 | 1.12 (0.72-1.72) |
| Model 3 | 1.00 (Referent) | **0.71 (0.54-0.94)** | 1.23 (0.94-1.61) | 0.81 (0.48-1.38) | 1.32 (0.59-2.94) | 0.641 | 1.00 (0.62-1.60) |
| Model 3 <sup>a</sup> | 1.00 (Referent) | 0.78 (0.56-1.09) | 1.15 (0.81-1.62) | 0.99 (0.55-1.80) | 1.77 (0.73-4.28) | 0.327 | 1.26 (0.75-2.13) |

Model 1: age (y), ethnic group (White, Asian/Asian British, Black/Black British, mixed), Townsend deprivation index, smoking (never, former, current [ $<10$ , $10-<50$ , $>50$ pack-years]), total physical activity (MET-h/week), diet habit, hypertension (yes, no), dyslipidemia (yes, no), diabetes (yes, no).
Model 2: Model 1 + BMI (kg/m<sup>2</sup>).
Model 3: Model 2 + other types of alcoholic beverages.
<sup>a</sup> Excluding participants who had reduced alcohol intake for illness or ill health, participants self-rating as having poor health, and participants with major cardiovascular disease or cancer at baseline, in addition to cases occurring within the first 2 years of follow-up.

Notably, in the exploratory analysis with multivariable adjustment (model 2), low (>0 to <1 serving/week) or moderate (2 to 3 servings/week) consumption of several specific alcoholic beverages was significantly associated with a lower risk of gout, including all the alcoholic beverages in men (**Table 4**) and red wine, champagne/white wine, and fortified wine in women (**Table 5**). All these inverse associations were attenuated to be null in the final analysis in which the specific alcoholic beverages were mutually adjusted for each other and potential reverse causation was further addressed. In particular, women with any levels of red wine intake had a lower risk of gout in the exploratory analysis (HR _per 1 glass/d_ = 0.79, 95% CI: 0.69-0.90), but not after adjusting for other alcoholic beverages and addressing potential reverse causation (HR _per 1 glass/d_ = 0.91, 95% CI: 0.77-1.06).

## Discussion

In a large population-based prospective study, we have assessed the consumption of total alcohol and several specific alcoholic beverages in relation to risk of gout. We first observed sex-specific associations between drinking status and risk of gout, with current drinkers having a higher risk than never drinkers in men but not in women. Among current drinkers, after addressing potential confounding and reverse-causation bias, greater alcohol consumption was associated with a substantially elevated risk of gout in men and a moderately elevated risk in women. Regardless of sex, greater consumption of several specific alcoholic beverages, especially beer/cider, was associated with a higher risk of gout.

In the majority of the prior studies, alcohol consumption was found to be associated with a higher risk of gout. A meta-analysis pooling data from 12 cohort studies revealed a combined relative risk of 1.83 when comparing the highest with non/occasional alcohol drinking (23).Very few previous studies of alcohol consumption and gout have included both men and women and assessed sex-specific associations. In a prospective analysis of a community-based Japanese cohort including 3,188 men and 6,346 women, higher amount of alcohol consumption (vs. never drinking) was associated with a higher risk of a composite outcome of hyperuricemia and gout in men, whereas no association was found in women (9). In the present study, we also found that male current drinkers (vs. never drinkers) had a substantially higher risk of gout. Female current drinkers showed a lower risk of gout than never drinkers after the multivariable adjustment, but this association was attenuated to be null after further adjusting for BMI or accounting for potential reverse causation.

In the additional analyses that were limited to current drinkers, we found that men who drank 5 times or more per week had an approximately 2-fold higher risk of gout than those who drank less than once per week. In female drinkers, there was initially an inverse association between drinking frequency and gout, but the association was reversed after further adjusting for BMI and especially after accounting for reverse-causation bias, with those drinking 5 times or more per week having a 29% higher risk of gout as compared with those drinking less than once per week. This sex-specific association may be, at least in part, owing to the difference in the type of alcohol consumed between men and women. In particular, women consumed considerably lower amount of beer/cider than men, which was most strongly associated with gout among the specific alcoholic beverages.

Only a few studies have assessed the consumption of specific alcohol and risk of gout and the findings are inconsistent. A prospective study of US male professionals (4) and the Framingham Heart Study (5) both found no correlation between wine consumption and gout, while in 2 other prospective studies (10, 11), wine consumption was associated with a higher risk of gout. In the current analysis of participants who self-identified as current drinkers, consumption of champagne/white wine, beer/cider, and spirits each was associated with a higher risk of gout in both sexes, with the strongest association observed for beer/cider, which is likely attributable to the greater content of purine in these alcoholic beverages (4).

We initially observed, both in male and female drinkers, that low to moderate consumption of several specific alcoholic beverages was associated with a lower risk of gout. After mutually adjusting for the specific alcoholic beverages and addressing potential reverse-causation bias, all these associations were nullified. It is likely that some participants with illness or ill health had abstained from alcohol before being recruited to the study, and self-identified as non-drinkers or occasional drinkers during the baseline assessment. Including such participants can increase the risk within the non-drinking or occasional drinking group, and lead to a U- or J-shaped association between alcohol consumption and health risk (15, 16). Our findings and other previous findings on alcohol (15, 16) and non-alcohol lifestyle factors (14) underscore the importance of considering reverse-causation bias for a more accurate estimation of the assessed relationship with health outcomes.

Our study possesses several strengths, including its large sample size, the prospective study design, the long-term follow up, and the comprehensive adjustment for potential confounders. To the best of our knowledge, the present study is among a few studies that have assessed the sex-specific relationships between the consumption of various alcoholic beverages and incident gout, and the first study that adequately addressed the potential reverse-causation bias.

The present study also has several potential limitations. Firstly, this is an observational study where the influence of residual confounding cannot be fully excluded. Secondly, because the frequency of alcohol consumption was self-reported, some degree of misclassification of exposure was inevitable. Thirdly, alcohol consumption was assessed at baseline and the influence of longitudinal changes in drinking pattern on gout remains to be investigated in future studies. Finally, the present study was based on the UK Biobank in which the majority of participants are of European descent who are ethnically white and are relatively healthier than the general population in the UK (24). Thus, further verification of the findings for other regional and ethnic populations is needed.

## Conclusions

In this prospective study with a careful consideration of potential confounding and reverse causation, consumption of total alcohol and several specific alcoholic beverages is associated with a higher risk of gout in both sexes. The observed sex-specific difference in the association of total alcohol consumption with incident gout is likely owing to difference in the type of alcohol consumed between men and women, rather than biological differences.

## Data Availability

The UK Biobank data are available upon application to the UK Biobank (www.ukbiobank.ac.uk/).

## Acknowledgements

The authors thank the UK Biobank participants. This research has been conducted using the UK Biobank Resource under Application Number 90087.

## Notes

### Competing Interest Statement

The authors have declared no competing interest.

### Funding Statement

This work is supported by the Gusu Leading Talent Plan for Scientific and Technological Innovation and Entrepreneurship (ZXL2023345).

### Author Declarations

The UK Biobank data are available upon application to the UK Biobank (www.ukbiobank.ac.uk/).

