## Supplementary materials for "Alcohol consumption and long-term risk of gout in men and women: a prospective study addressing potential reverse causation"

**Supplementary Table 1. Baseline characteristics of current drinkers in men and women according to drinking frequency**

|  | **Drinking frequency in men** | | | | **Drinking frequency in women** | | | |
| --- | --- | --- | --- | --- | --- | --- | --- | --- |
|  | **<1/week** | **1-2/week** | **3-4/week** | **>=5/week** | **<1/week** | **1-2/week** | **3-4/week** | **>=5/week** |
| **Participants, n** | 36,201 | 56,976 | 56,868 | 54,614 | 76,286 | 69,913 | 55,719 | 43,692 |
| **Age at baseline, y** | 55.86 ± 8.59 | 55.89 ± 8.39 | 56.54 ± 8.04 | 58.06 ± 7.66 | 56.27 ± 8.18 | 55.77 ± 8.03 | 55.82 ± 7.83 | 57.39 ± 7.61 |
| **Ethnicity, %** |  |  |  |  |  |  |  |  |
| White | 92.85 | 96.51 | 97.84 | 98.14 | 94.35 | 97.72 | 98.56 | 98.77 |
| Asian or Asian British | 3.32 | 1.57 | 1.03 | 0.92 | 1.76 | 0.60 | 0.36 | 0.30 |
| Black or Black British | 3.08 | 1.43 | 0.77 | 0.57 | 2.97 | 1.10 | 0.59 | 0.42 |
| Mixed | 0.75 | 0.49 | 0.37 | 0.37 | 0.92 | 0.58 | 0.49 | 0.51 |
| **Townsend deprivation index** | -0.76 ± 3.37 | -1.31 ± 3.10 | -1.68 ± 2.91 | -1.55 ± 2.97 | -0.91 ± 3.20 | -1.55 ± 2.92 | -1.84 ± 2.73 | -1.71 ± 2.80 |
| **Smoking status, %** |  |  |  |  |  |  |  |  |
| Never | 54.85 | 54.24 | 50.24 | 37.90 | 63.58 | 62.42 | 56.96 | 45.33 |
| Former | 30.81 | 34.48 | 39.56 | 46.79 | 26.48 | 29.74 | 35.81 | 43.43 |
| Current: <10 pack-years | 1.42 | 1.15 | 1.20 | 1.94 | 1.18 | 1.10 | 1.28 | 1.94 |
| Current: 10-50 pack-years | 11.27 | 9.21 | 8.22 | 11.51 | 8.21 | 6.44 | 5.69 | 8.61 |
| Current: >50 pack-years | 1.65 | 0.92 | 0.78 | 1.87 | 0.56 | 0.31 | 0.27 | 0.69 |
| **Physical activity, MET-h/week** | 44.97 ± 48.05 | 45.70 ± 46.57 | 43.45 ± 42.94 | 43.35 ± 44.46 | 38.77 ± 37.67 | 38.78 ± 35.83 | 38.68 ± 35.00 | 39.53 ± 36.68 |
| **Diet score** | 2.42 ± 1.40 | 2.39 ± 1.38 | 2.43 ± 1.36 | 2.31 ± 1.33 | 2.95 ± 1.33 | 2.95 ± 1.30 | 2.97 ± 1.28 | 2.89 ± 1.27 |
| **Hypertension, %** | 60.78 | 60.53 | 62.59 | 67.42 | 50.90 | 47.44 | 47.12 | 51.55 |
| **Dyslipidemia, %** | 25.92 | 22.71 | 22.40 | 24.50 | 16.54 | 12.45 | 10.69 | 11.71 |
| **Diabetes, %** | 11.44 | 7.63 | 5.50 | 5.61 | 6.34 | 3.26 | 2.14 | 2.11 |
| **BMI, kg/m^2^** | 28.29 ± 4.78 | 27.98 ± 4.22 | 27.53 ± 3.84 | 27.25 ± 3.76 | 28.19 ± 5.76 | 26.97 ± 4.89 | 26.15 ± 4.43 | 25.74 ± 4.25 |
| **Average consumption of specific alcoholic beverages** | | | |  |  |  |  |  |
| Red wine ^a^ | 0.14 ± 0.22 | 1.63 ± 2.26 | 3.99 ± 4.15 | 5.87 ± 5.58 | 0.12 ± 0.21 | 1.55 ± 1.95 | 3.35 ± 3.27 | 4.34 ± 4.26 |
| Champagne/white wine ^a^ | 0.12 ± 0.20 | 0.81 ± 1.51 | 1.68 ± 2.56 | 2.35 ± 3.51 | 0.16 ± 0.22 | 1.61 ± 2.02 | 3.13 ± 3.35 | 4.11 ± 4.43 |
| Beer/cider ^b^ | 0.21 ± 0.24 | 3.17 ± 3.31 | 4.82 ± 4.85 | 5.32 ± 5.76 | 0.06 ± 0.15 | 0.46 ± 1.08 | 0.53 ± 1.22 | 0.51 ± 1.30 |
| Spirit ^c^ | 0.75 ± 0.17 | 0.97 ± 2.37 | 1.64 ± 3.32 | 2.84 ± 5.16 | 0.08 ± 0.17 | 0.92 ± 1.90 | 1.20 ± 2.36 | 1.56 ± 3.07 |
| Fortified wine^a^ | 0.02 ± 0.09 | 0.10 ± 0.48 | 0.16 ± 0.65 | 0.24 ± 0.9 | 0.03 ± 0.10 | 0.17 ± 0.62 | 0.25 ± 0.84 | 0.32 ± 1.09 |

BMI, body mass index; MET, metabolic equivalent.

^a^ glasses/week, ^b^ pints/week, ^c^ measures/week.

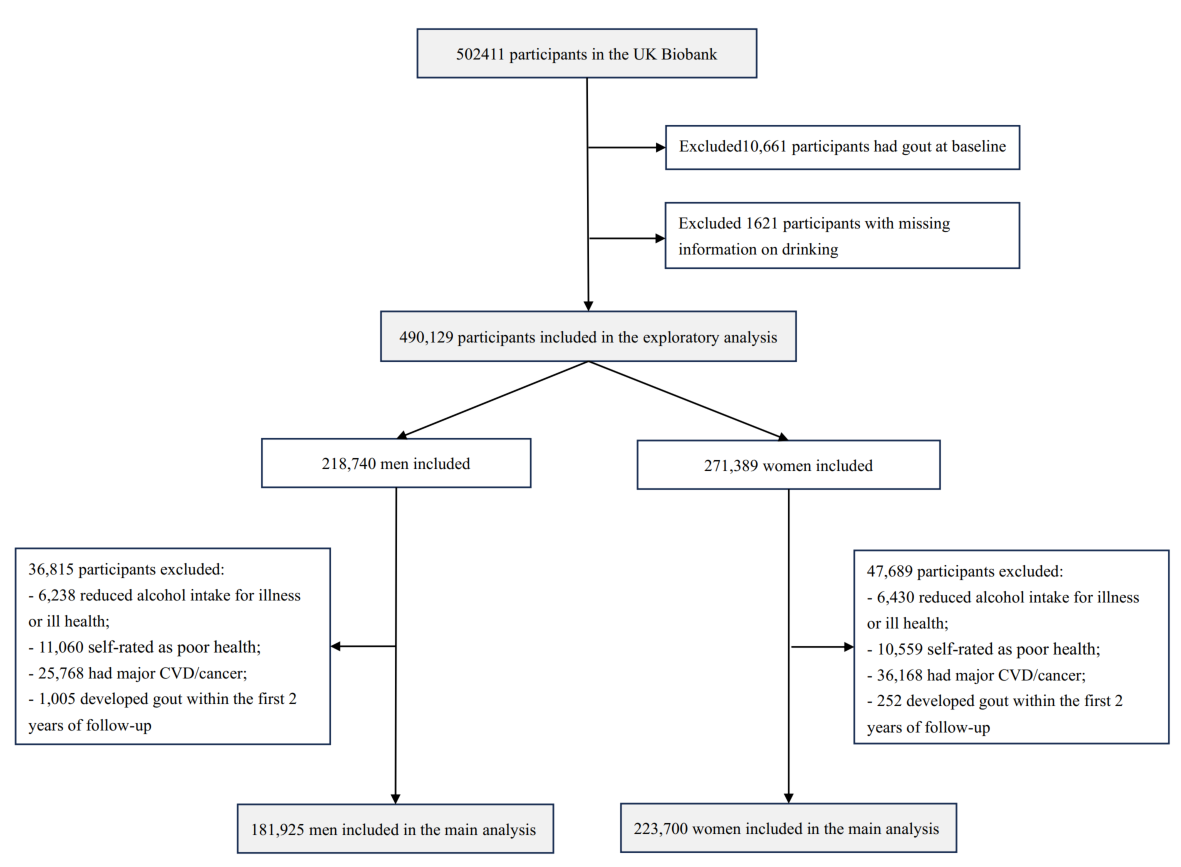

**Supplementary Figure 1. Flow diagram of participant selection**

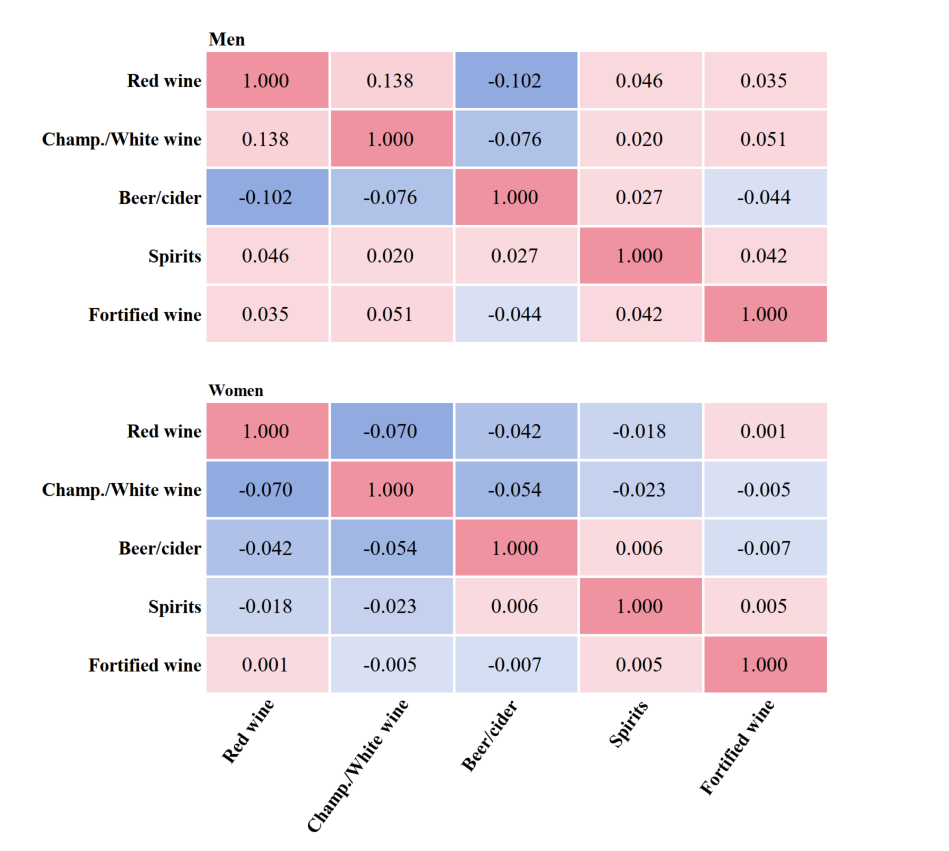

**Supplementary Figure 2.** **Spearman partial correlations among specific alcoholic beverages in men and women**

Results were adjusted for age (y), ethnic group (White, Asian/Asian British, Black/Black British, mixed), and Townsend deprivation index.
